# Retention in care and viral suppression among people on HIV treatment receiving 6- versus 12-month prescriptions in South Africa: a cohort study using routinely collected electronic health data

**DOI:** 10.64898/2026.11.25.26368936

**Authors:** Oratile Mokgethi, Amy Huber, Khumbo Shumba, Maggie Munsamy, Nishana Ramdas, Lise Jamieson, Sydney Rosen, Merlin Pillay, Sophie Pascoe, Musa Manganye, Lufuno Malala, Matthew P. Fox, Jacqui Miot, Idah Mokhele

## Abstract

**Introduction:** During the COVID-19 pandemic, ART prescription lengths in South Africa were temporarily extended from six months to twelve months for stable clients, reducing required clinic visits from two to one per year. We compared retention and viral suppression outcomes between clients receiving 6-month and 12-month prescriptions within South Africa’s Centralised Chronic Medicines Dispensing and Distribution (CCMDD) programme, which serves over 3.2 million people living with HIV through medication collection at facility-based or community-based pickup points.

**Methods:** We conducted a retrospective cohort study in seven South African provinces using routinely collected electronic health data. We included adults (≥18) with a 6-month or 12-month ART prescription issued between June and November 2020. Participants were followed for 24 months. Retention was defined as ≥1 medication collection visit during months 13-24 of follow-up and viral suppression as a viral load <50 copies/ml at the first test conducted in that window. We estimated unadjusted retention and suppression rates and then used a modified Poisson regression model with a log link and robust standard errors to compare non-retention and viral non-suppression between groups, accounting for district-level clustering and adjusting for age, sex, time on ART, model of care, and ART regimen.

**Results:** Among 428,926 clients, 196,531 (46%) received 6-month prescriptions and 232,395 (54%) received 12-month prescriptions. During the outcome assessment period (months 13 to 24), clients in the 6-month group attended a median of 2 clinical prescribing visits and clients in the 12-month group attended a median of 1 clinical prescribing visit. Unadjusted retention was 91.4% in the 6-month group and 91.6% in the 12-month group; viral suppression was 79.3% in the 6-month group and 80.9% in the 12-month group. After adjustment, using the 6-month prescription group as the reference, adjusted risk ratios were 0.98 (0.91–1.05) for non-retention and 0.93 (0.82–1.06) for viral non-suppression.

**Conclusion:** Twelve-month prescriptions maintained retention in care and viral suppression rates equal to 6-month prescriptions while halving required annual clinic visits per client. These findings support further evaluation of reduced-frequency prescribing within differentiated HIV service delivery programmes for stable clients and contribute to the evidence base to guide recommendations on reduced clinical prescribing visits.

## Introduction

In South Africa, where some 6.2 million people are receiving antiretroviral therapy (ART) in the public health sector(1), ART clients are traditionally issued six-month prescriptions(2,3) requiring two full clinic visits per year with interim 2- or 3-month medication refills. The South African National Department of Health (NDOH) introduced facility-based six multi-month dispensing (6MMD) in 2025, with national rollout in 2026, allowing eligible clients to obtain all their medications at their six-monthly clinic visits but still requiring two full clinics visit per year(4).

During the COVID-19 pandemic, the South African Government temporarily extended prescription validity to 12 months, reducing required clinic visits to one per year while maintaining regular 2-3-month medication pickups(5). During this period, both 6- and 12-month prescriptions were issued, an opportunity to compare of 6- and 12-month scripting in routine care and to estimate the effect of scripting duration on clinical outcomes for which there is currently little evidence(6). The World Health Organization (WHO), which currently recommends 6-monthly clinic visit frequency for clients established on ART, has called for data on this question(2), as understanding whether 12-month prescriptions maintain comparable outcomes to the standard 6-month approach is crucial for informing future policy decisions and optimizing DSD models in resource-constrained settings.

This study evaluates outcomes among stable HIV clients enrolled in differentiated models of care (DMOC) in South Africa, comparing those with 6- and 12-month prescription lengths and providing essential data to guide the design of sustainable, patient-centered HIV care models in the post-pandemic era.

## Methods

### Study context

We conducted a cohort analysis using routinely collected, de-identified electronic data from Tier.Net and SYNCH (Synchronized National Communication in Health) in public health sector clinics acoss seven of South Africa’s nine provinces (Eastern Cape, Free State, Gauteng, KwaZulu-Natal, Limpopo, North West, and Mpumalanga). Western Cape was excluded because it operates a different data infrastructure and does not use one of the study databases (SyNCH); Northern Cape was excluded because the province had not yet fully adopted SyNCH during the study period and was still largely relying on paper-based prescription records.

At the time of this study (2020–2022), South African treatment guidelines for HIV treatment monitoring called for viral load testing at 6 and 12 months after ART initiation, followed by annual testing(7). Stable clients, defined as those who had been on ART for ≥ 6 months, were virally suppressed (<50copies/ml), and did not have active tuberculosis (TB), pregnancy or uncontrolled chronic conditions, were eligible for enrollment in one of South Africa’s differentiated models of care (DMOCs, also referred to as differentiated service delivery or DSD models(8)). The two most widely used DMOCs were facility-based fast-track medication pickup (Fac-PuP) and community-based medication pickup (Ex-PuP)(9,10), both using medications distributed by the Centralised Chronic Medicines Dispensing and Distribution (CCMDD) program.

Under the CCMDD models, eligible clients receive a 6-month prescription at a medical visit with a clinician, where they are dispensed two or three months of medication (standard dispensing intervals were extended from two to three months in May 2023(11)). Interim medication supplies between clinic visits are collected from facility-based pickup points or community-based external pickup point locations without queuing or seeing a clinician.

Between May and December 2020, prescription validity was temporarily extended to 12 months for stable clients. Clients continued to collect medication every 2-3 months from their usual pickup points, but full clinic visits previously required twice a year were reduced to once per year for those issued 12-month prescriptions. The 12-month option was offered at the discretion of the prescribing clinician and may have reflected perceptions of client reliability or need, medication stock availability, and/or clinician preference.

### Study population

The study population included adults (aged 18+ years) on ART who received either a 6-month or a 12-month prescription visit between 1 June and 30 November 2020. We excluded clients who did not have a prescribing visit within the baseline period, clients younger than 18 years at baseline, or were from provinces not included in the study (Northern Cape and Western Cape).

### Data sources and data management

We analysed data from two systems: 1) de-identified client data from TIER.Net, an electronic patient management system that tracks HIV/ART client information including demographic, clinical and clinic visit data for all clients on ART in the South African public health sector, excluding the Western Cape(12); and 2) de-identified client data from the SyNCH(13), which tracks CCMDD programme activities including clinical prescribing visits, at which a clinician issues a prescription, and medication collection visits, at which clients collect their dispensed medication from a facility-based or community-based pickup point without seeing a clinician.

Data from SyNCH and TIER.Net were linked by the National Department of Health and de-identified before being provided for analysis. Records were matched sequentially, using the Health Patient Registration System (HPRS) number as the primary linkage variable, supplemented by national identification number, name, and date of birth where HPRS was unavailable. For this analysis, we included only perfect matches, defined as exact agreement on at least one unique identifier (HPRS number or national identification number), confirmed by patient name and surname and facility name.

### Variables and outcomes

We defined time zero (i.e. the baseline date) as the date of the first prescription issued during the enrolment period (1 June to 30 November 2020), which captured the interval during which prescriptions could be either 6 or 12 months in length. The exposure variable was defined as the prescription length, either 6 or 12 months, recorded at the baseline visit. Where a client received a 6-month prescription but transitioned to a 12-month prescription at their subsequent visit, we retained the baseline prescription length as the exposure. Baseline variables included each participant’s DMOC (Fac-PuP or Ex-PuP), ART regimen, and comorbidity status. ART regimen was categorised as first line TLD (tenofovir/lamivudine/dolutegravir, TDF/3TC/DTG), first line TEE (tenofovir/emtricitabine/efavirenz, TDF/FTC/EFV), or other. Where regimen data were incomplete, the first non-missing ART regimen after the baseline visit was assumed. Comorbidity status was classified as HIV without a recorded comorbid condition or HIV with one or more non-HIV comorbid conditions, identified through relevant ICD-10 (International Classification of Diseases, 10th Revision) codes in the client’s record and prescription of condition-specific medications.

Retention in care and viral suppression, assessed between months 13 and 24 of follow-up after the baseline visit, were the primary outcomes for this analysis. Clients were considered retained if they had at least one medication dispensing or collection visit during the outcome assessment window (months 13-24) (Fig. 1). Clients were considered not retained if they had no medication dispensing or collection visit during this period. Transfers to other facilities and deaths were included in the definition of those not retained.

**Figure 1:**
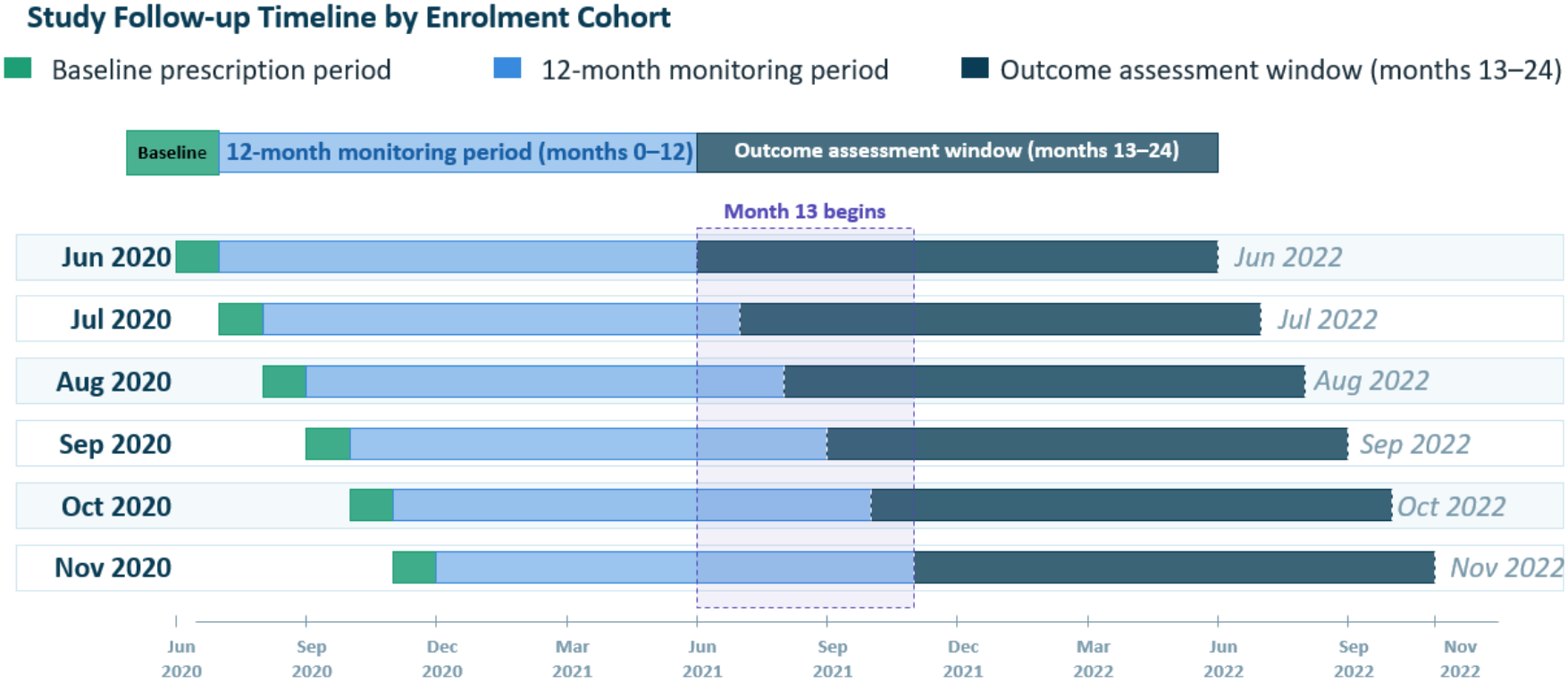
Illustration of study endpoints by enrolment cohort.

Viral suppression was defined as a viral load <50 copies/ml on the first test recorded in months 13-24, consistent with South African guidelines(4). Clients without a documented viral load measurement were excluded from the primary viral suppression analysis, though included in sensitivity analysis, as we could not ascertain whether missing viral load results reflected true disengagement from care or simply a missed laboratory test during a period of reduced health system capacity. In sensitivity analysis, clients with missing viral load results were instead classified as either all virally unsuppressed or all virally suppressed.

### Statistical analysis

Baseline characteristics were summarised using median and interquartile range (IQR) for continuous variables and frequencies with percentages for categorical variables. Relative risks of retention in care and viral suppression were estimated with a modified Poisson regression model with a log link and robust standard errors. The model accounted for clustering across 41 districts at the district level, as facilities within the same district tend to share administrative and operational characteristics that make their outcomes more similar to each other than to facilities elsewhere. We chose to cluster at the district rather than the facility level because the large number of facilities relative to the number of clients per facility would have made facility-level clustering unstable, and district-level clustering provided a more reliable way to account for this correlation. Covariates selected a *priori* based on relevance to viral suppression and retention in care included age, sex, time on ART at baseline, model of care, ART regimen at baseline, and presence of co-morbid conditions. We excluded clients with missing or unclassified values for sex, DSD model type, or ART regimen because numbers were small. Characteristics were also disaggregated by comorbidity. The potential impact of unmeasured confounding arising from the non-random assignment of prescription length was calculated with an E-value for the primary exposure association, quantifying the minimum strength of association that an unmeasured confounder would need to have with both the exposure and the outcome to fully explain the observed association.

### Artificial intelligence (AI) disclosure statement

During the preparation of this work, Claude Sonnet 4.6 was used to generate figures from tables provided by the authors. ChatGPT 5.6 Sol was used to check internal consistency of data and results reported, edit text for clarity, and copyedit text. The authors conducted all analysis and take full responsibility for the accuracy, integrity, and final content of this publication.

## Results

### Study sample

From the initial total of 3,168,298 linked records, this resulted in 1,068,135 perfect matches for our analysis; the remainder were partial matches that did not meet the minimum linkage threshold. Of these, 430,735 had a clinical prescribing visit recorded during the baseline period of June to November 2020. After applying eligibility criteria, the final study sample included 428,926 clients across seven provinces. Of these, 196,531(46%) received a 6-month prescription and 232,395 (54%) received a 12-month prescription at baseline (Figure 2).

**Figure 2.**
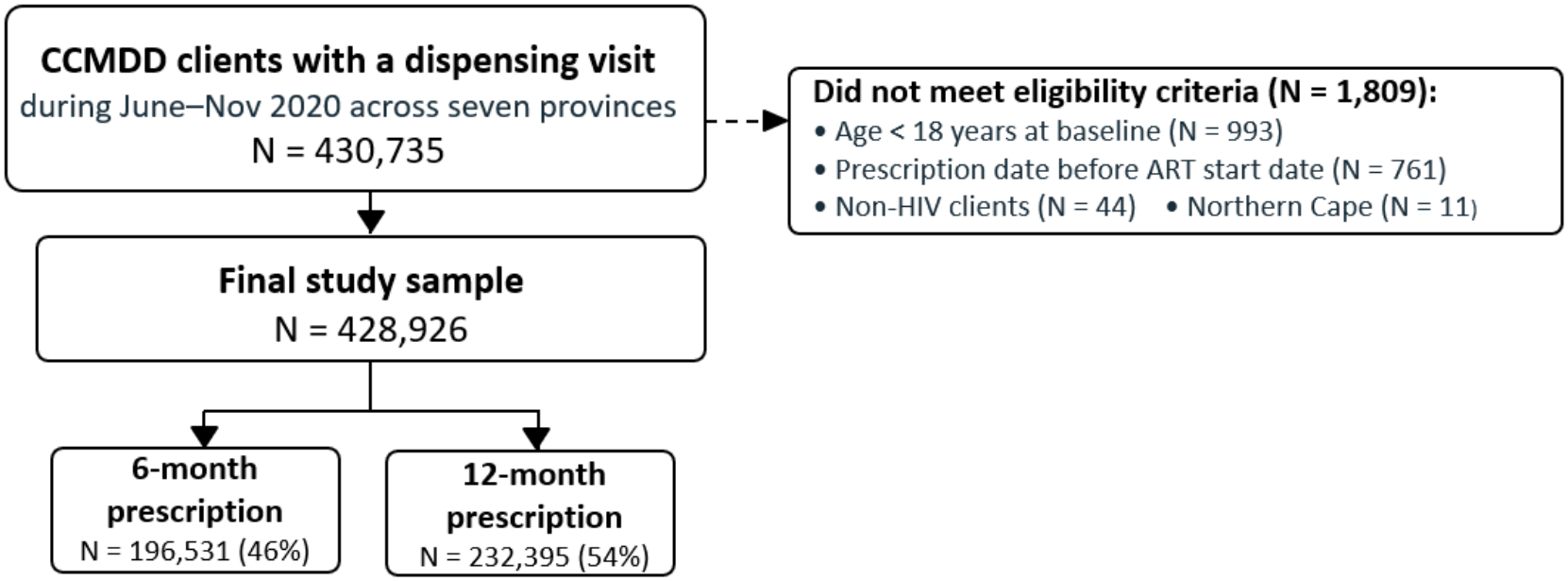
Study sample and allocation to prescription length groups.

### Participant characteristics in each prescription duration group

Of the 428,926 clients included in the analysis, 46% received 6-month prescriptions and 54% received 12-month prescriptions, as shown in Table 1. The two groups were broadly similar across most baseline characteristics. The majority of clients in both groups were female, at 67% and 69% respectively, with a median age of 43-44 years, and most had been on ART for five to nine years. The distribution among models of care and provinces was similar across both prescription groups. The proportion of clients on first-time TLD was higher in the 12-month group (60%), however, than in the 6-month group (49%), reflecting the national rollout of dolutegravir-based regimens during the study period. Of the 196,530 clients assigned to the 6-month prescription group at baseline, 80,160 (40.8%) received at least one 12-month prescription during follow-up, transitioning at a median of 7.4 months from baseline (IQR 5.6–11.0).

**Table 1:** Demographic and clinical characteristics of study participants by first prescription length, CCMDD programme, South Africa, June–November 2020 (n=428,926)

| **Characteristic (N, row %)** | **6-month prescription** | **12-month prescription** |
|---|---|---|
| N | 196,531 (46) | 232,395 (54) |
| Sex (female) | 131,785 (45) | 160,459 (55) |
| Age in years |  |  |
| Median (Q1, Q3) | 44 (37, 51) | 43 (37, 51) |
| 18–24 | 5,488 (42) | 7,495 (56) |
| 25–34 | 42,237 (44) | 54,580 (56) |
| 35–49 | 100,155 (46) | 117,057 (54) |
| 50+ | 38,075 (47) | 42,602 (53) |
| Missing | 10,576(50) | 10,661(50) |
| Time on ART at baseline |  |  |
| Median years (Q1, Q3) | 6 (4, 9) | 6 (4, 9) |
| 6 months–1 year | 9,577 (42) | 13,273 (58) |
| 2–4 years | 58,838 (45) | 72,033 (55) |
| 5–9 years | 92,195 (47) | 105,837 (53) |
| 10+ years | 35,921 (47) | 41,252 (53) |
| Model of care |  |  |
| Ex-PuP (external\community pickup up point) | 122,600 (45) | 148,047 (55) |
| Fac-PuP (facility based pickup point) | 73,597 (47) | 84,284 (53) |
| Missing | 334 (84) | 64 (16) |
| ART regimen |  |  |
| First-line TLD | 95,848 (41) | 139,693 (59) |
| First-line TEE | 92,146 (53) | 82,166 (47) |
| Other | 8,493 (45) | 10,469 (55) |
| Missing | 44 (39) | 67 (61) |
| Province |  |  |
| Eastern Cape | 20,423 (42) | 27,853 (58) |
| Free State | 5,983 (36) | 10,613 (64) |
| Gauteng | 53,066 (48) | 56,706 (52) |
| KwaZulu-Natal | 79,291 (45) | 96,188 (55) |
| Limpopo | 2,150 (29) | 5,330 (71) |
| Mpumalanga | 24,609 (51) | 23,410 (49) |
| North West | 11,009 (47) | 12,295 (53) |
| Numbers of visits during months 13–24 (median, IQR) |  |  |
| Clinical prescribing visits | 2 (1,2) | 1 (1,2) |
| Medication collection visits | 3 (2,4) | 3 (2,4) |
| Total interactions | 3(2,5) | 3(2,5) |

*ART, antiretroviral therapy; TLD, tenofovir disoproxil fumarate/lamivudine/dolutegravir; TEE, tenofovir disoproxil fumarate/emtricitabine/efavirenz; Ex-PuP, external/community pickup point; Fac-PuP, facility-based pickup point; CCMDD, Centrally Chronic Medicines Dispensing and Distribution; IQR, interquartile range; Q1, first quartile; Q3, third quartile. All percentages are row percentages showing the distribution between 6-month and 12-month.*

Clients with 12-month prescriptions made a median of 1 clinical prescribing visit during the outcome assessment period (mean 1.4), compared to a median of 2 (mean 1.6) for clients with 6-month prescriptions. Medication collection visits were similar between groups (median 3, mean 2.7 vs 2.9).

### Retention outcomes by prescription length

The overall proportion of clients prescribed 12-month scripts peaked in August 2020 at 63%, before declining through November 2020 (Figure 3a). Prescription of 12-month scripts varied across provinces, with Limpopo and Free State having the highest proportions and Mpumalanga the lowest (Figure 3b).

**Figure 3.**
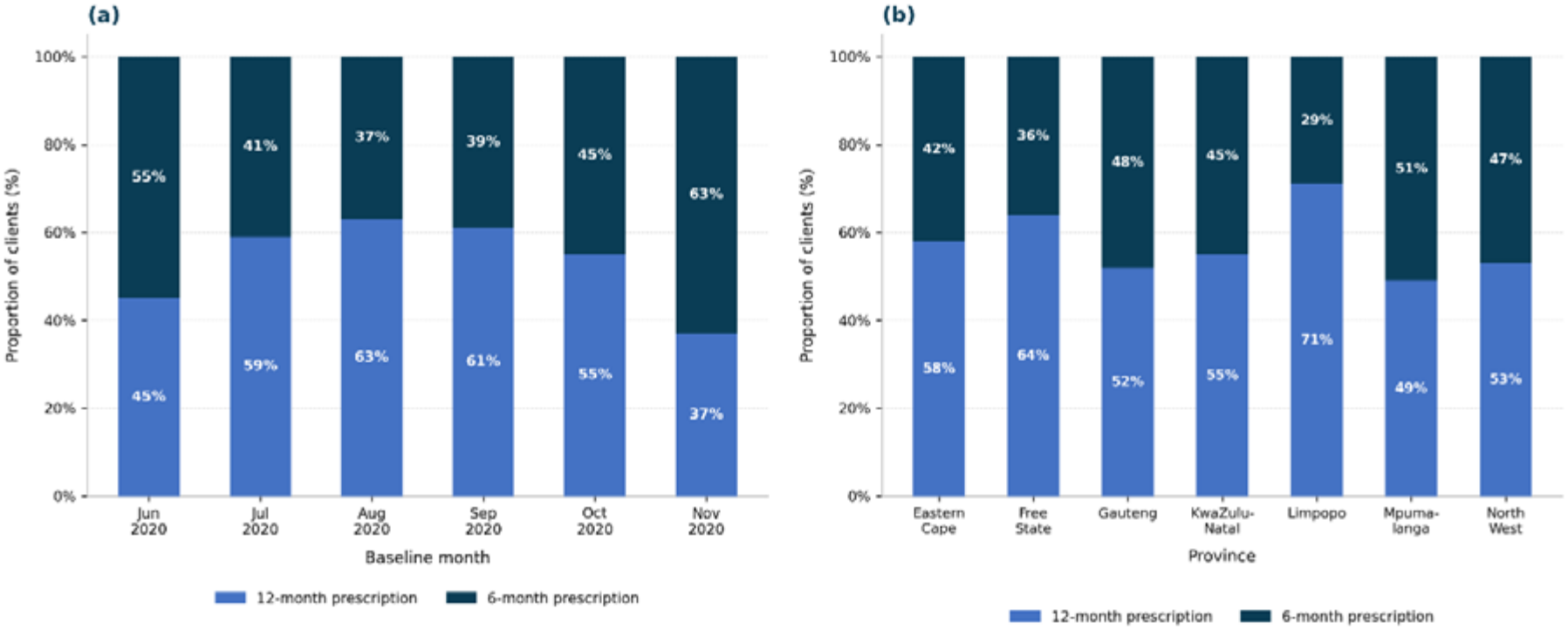
Distribution of first prescription length among CCMDD programme clients, by baseline enrolment month (a) and province (b), South Africa, June–November 2020 (N = 428,926).

Retention in care during the 13–24-month period exceeded 90% in both 6- and 12-month prescription groups in every province, ranging from 90.9% to 92.0% for 6-month prescriptions and from 90.9% to 92.1% for 12-month prescriptions (Figure 4). Gauteng had the highest retention in both groups at 92.0% and 92.1% respectively, while Eastern Cape had the lowest at 90.9% in both groups. Observed differences in retention between the two prescription groups in any province was too small to be meaningful after adjustment. Nearly half (40.8%) of clients in our 6-month group switched to 12-month prescriptions during the follow-up period but remained in the 6-month group for analysis, suggesting that the differences we observed between groups underestimate the true effect of 12-month prescribing. A formal interaction test confirmed no significant difference in the prescription length effect during June–September 2020, though the effect was more pronounced among clients enrolled in October and November 2020.

**Figure 4:**
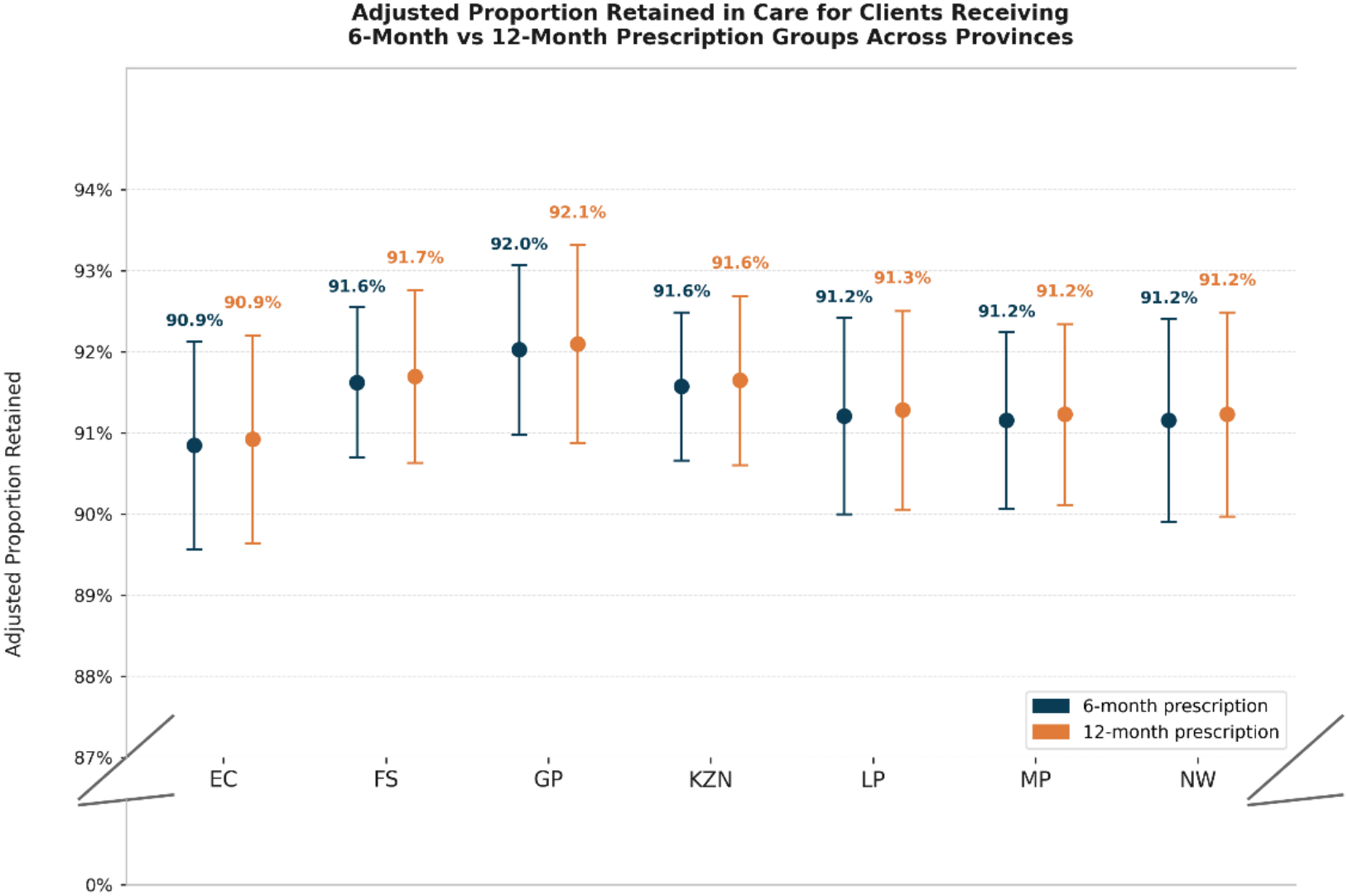
Adjusted proportion of clients retained in care during months 0–12, by first prescription length and province, South Africa, June–November 2020 (N = 428,926).

After adjustment, clients in the 12-month prescription group had no difference in non-retention compared to those in the 6-month group (aRR 0.98, 95% CI 0.91–1.05), confirming that extending prescription length was not associated with poorer retention in care (Figure 5, Supplementary table S2).

**Figure 5.**
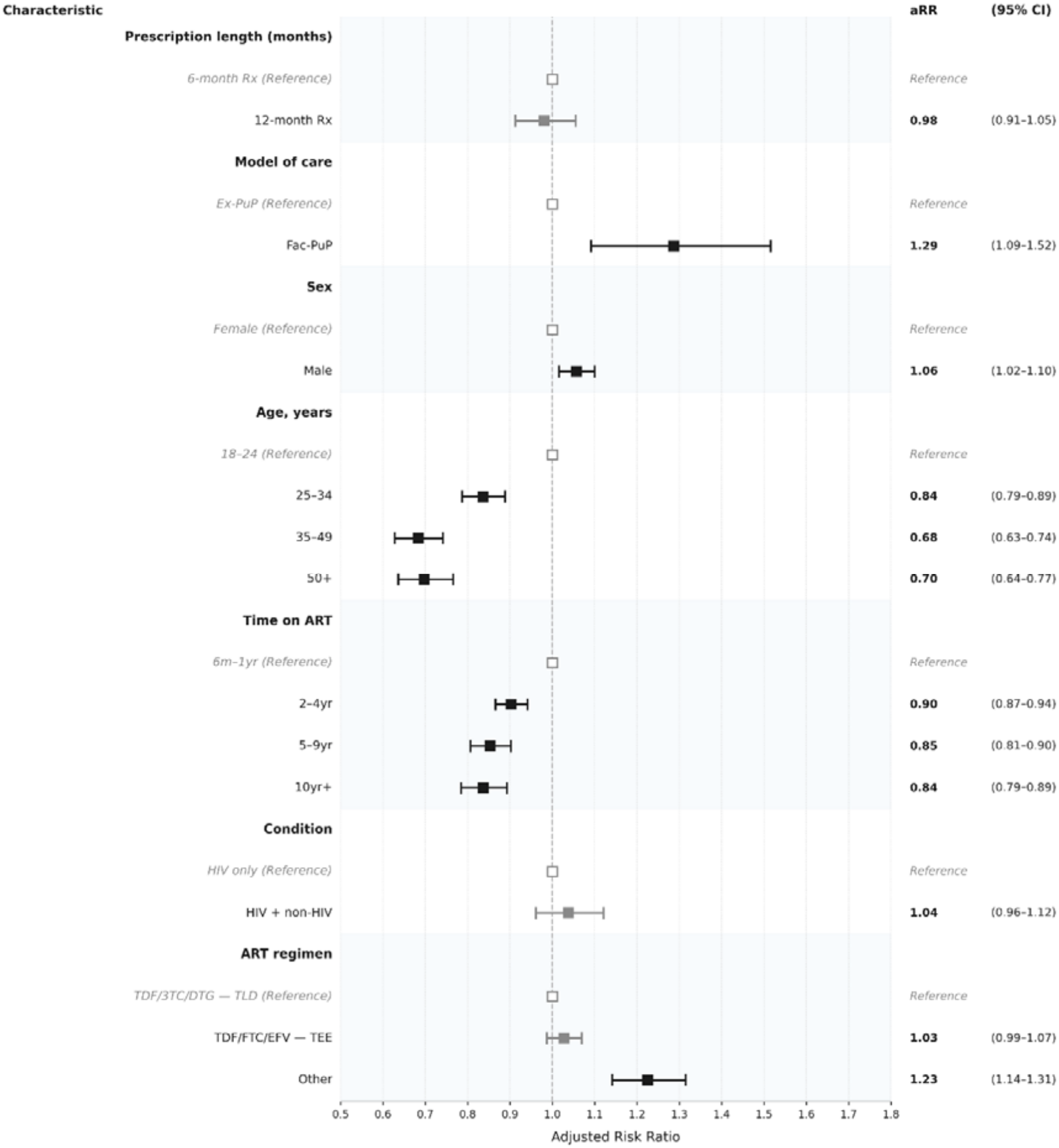
Adjusted relative risk of non-retention in care by baseline characteristics among people living with HIV receiving 6-month versus 12-month prescriptions in South Africa’s CCMDD programme, June–November 2020 (N = 428,926) *aRR = adjusted risk ratio. CI = confidence interval. Outcome = non-retention in care. Adjusted for: prescription length, model of care, sex, age, time on ART, condition, ART regimen, and province. Filled squares = statistically significant. Open squares = reference categories*.

Other associations with non-retention were small. Clients collecting medication from a facility pickup point were more likely to be not retained compared to those using community-based external pickup points (aRR 1.29, 95% CI 1.09–1.52). Female clients, older clients, and those who had been on ART for longer were more likely to be retained. Comorbidity status was not associated with non-retention after adjustment. Unsurprisingly, clients in the other ART regimen category, which comprises regimens prescribed following confirmed virological failure on a first-line regimen, showed higher non-retention compared to TDF/3TC/DTG (aRR 1.23, 95% CI 1.14–1.32).

### Viral suppression status by prescription length

Across all seven provinces, viral suppression ranged from 78.6% to 79.4% in the 6-month group and from 79.8% to 80.7% in the 12-month group. Viral load results were available for 79% of clients in the 6-month group (154,311 of 196,531) and 71% in the 12-month group (165,008 of 232,395), indicating that a higher proportion of clients in the 12-month group did not have a documented viral load result. Although viral suppression was numerically higher in the 12-month group in every province, the overall adjusted association showed no difference (Figure 6).

**Figure 6:**
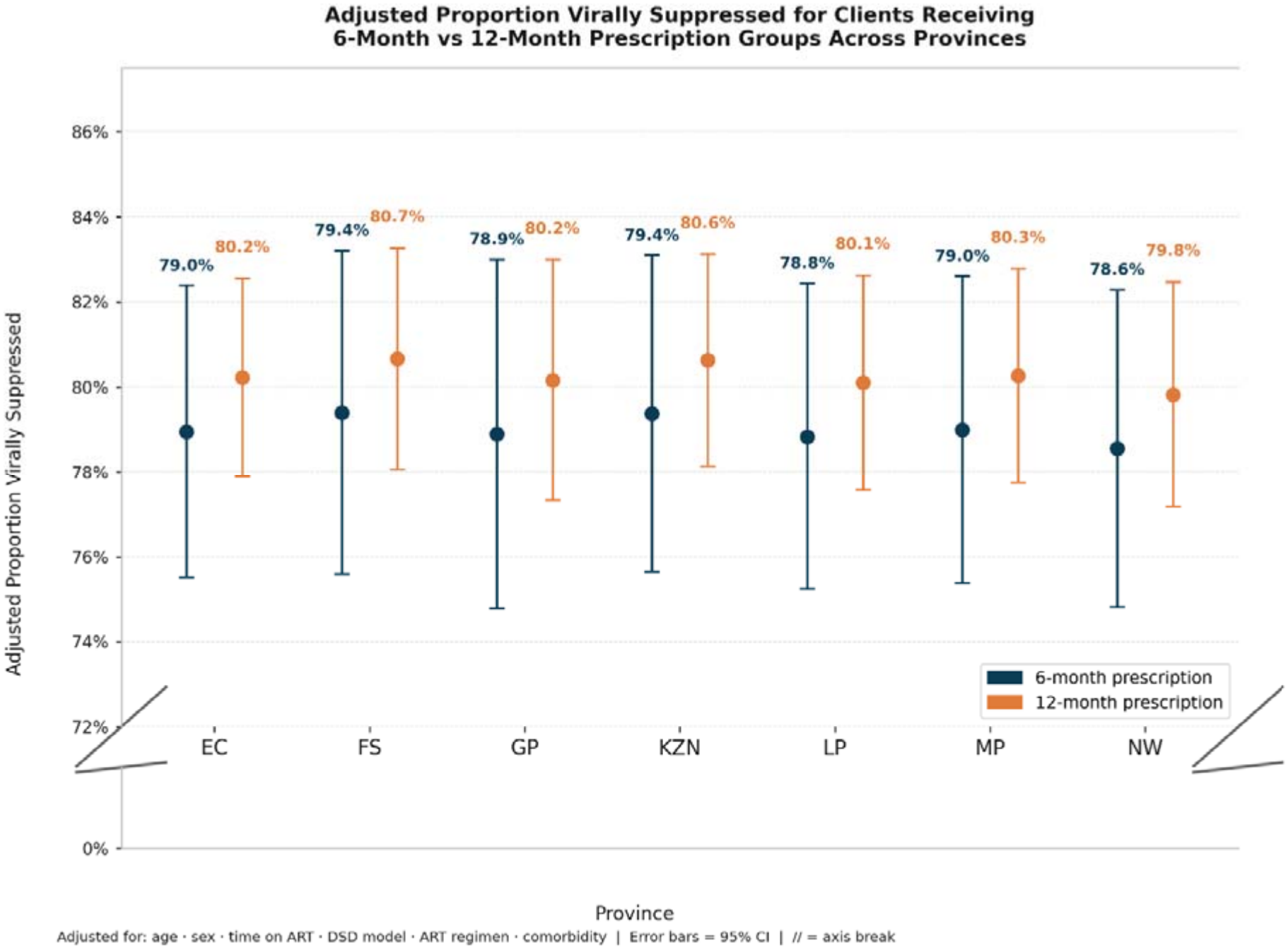
Adjusted proportion of clients with viral suppression at months 13–24, by first prescription length and province, South Africa, June–November 2020.

**Figure 7.**
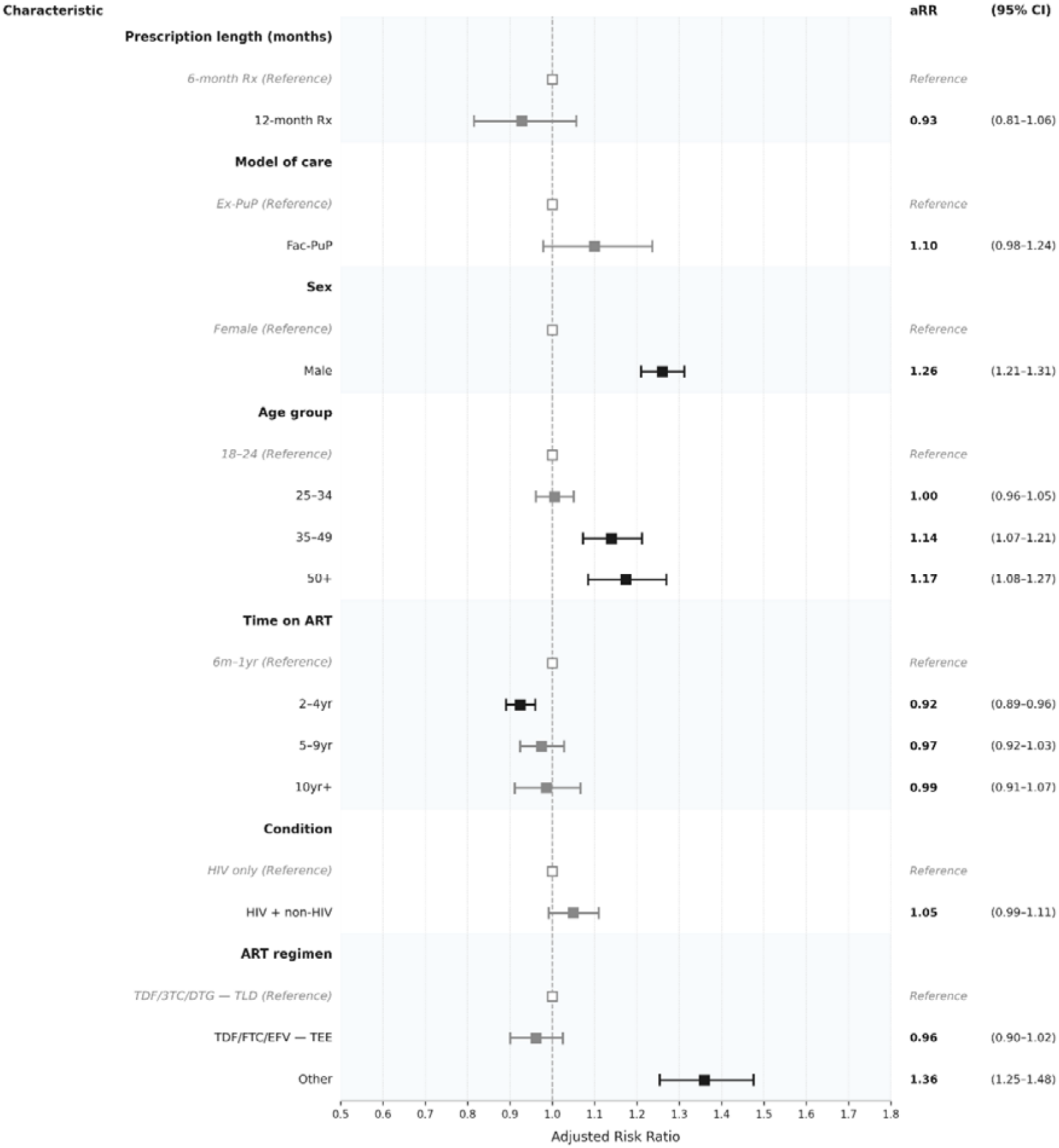
Adjusted relative risk of not achieving viral suppression by baseline characteristics among people living with HIV receiving 6-month versus 12-month prescriptions in South Africa’s CCMDD programme, June–November 2020 (N = 319,319 retained clients with a recorded viral load result) *aRR = adjusted risk ratio. CI = confidence interval. Outcome = not achieving viral suppression (viral load ≥50 copies/ml). Adjusted for: prescription length, model of care, sex, age group, time on ART, condition, ART regimen, and province. Filled squares = statistically significant. Open squares = reference categories*.

Of the 392,488 clients retained in care, 319,319 had a recorded viral load result during the outcome assessment period. Of these, 255,755 (80%) were virally suppressed. After adjustment, clients in the 12-month prescription group were not more likely to be virally unsuppressed than those in the 6-month group (aRR 0.3, 95% CI 0.81–1.06). Model of care was not associated with viral non-suppression after adjustment.

Male clients were significantly more likely to be virally unsuppressed than female clients (aRR 1.26, 95% CI 1.21–1.31), one of the strongest associations in the model. Older age was also associated with being virally unsuppressed. Compared to clients aged 18 to 24 (and 25-35, who were not different from the youngest age group), those aged 35 and above were significantly more likely to be virally unsuppressed. Clients who had been on treatment for two to four years were slightly less likely to be virally unsuppressed than those in their first year (aRR 0.92, 95% CI 0.89–0.96), though this protective association was not observed among clients on treatment for five years or more. Clients on regimens other than TDF/3TC/DTG were significantly more likely to be virally unsuppressed (aRR 1.36, 95% CI 1.25–1.48), while comorbidity status was not associated with being virally unsuppressed after adjustment (Supplementary Table S3).

### Sensitivity analysis

When clients with missing viral load results were reclassified as virally unsuppressed in a sensitivity analysis, the prescription length finding changed direction (Supplementary Table S4): clients in the 12-month group were significantly more likely to not achieve viral suppression than those in the 6-month group (aRR 1.15, 95% CI 1.10–1.20), while the proportion classified as virally suppressed remained higher in the 6-month group. Findings for all other covariates remained broadly consistent in direction with the primary analysis (Supplementary Table S4).

## Discussion

In this large retrospective cohort of 428,926 people living with HIV in seven of nine South African provinces, we found high overall rates of retention in care and viral suppression among clients receiving both 6-month and 12-month CCMDD prescriptions. Adjusted retention in care was consistently above 90% across all provinces, with no meaningful difference (aRR 0.98, 95% CI 0.91– 1.05) between clients receiving 6-month and 12-month prescriptions. There was no difference in viral non-suppression between prescription groups once adjusted for demographic and clinical characteristics. In this cohort, extending prescription length to 12 months did not compromise either outcome and resulted in a halving of clinic visits, an important benefit for both clients and the health system.

Retention in care was high in this study, consistent with an earlier single-province South African study reporting that among 27,148 clients referred for community ART delivery in KwaZulu-Natal, retention in care was 94.6% among clients receiving 12-month scripts and 91.8% among those receiving 6-month scripts (14). Also, largely consistent with the limited existing evidence, viral suppression was also high across both our groups, with no difference between clients receiving 6- and 12-month prescriptions in adjusted analysis. Studies in Zambia, Zimbabwe, and Uganda similarly found that extending prescription intervals does not compromise viral suppression(15–17). In a systematic review (6) comparing 6- and 12-monthly clinic visits to standard 3 monthly visits--a slightly different comparison than the 6 versus 12-month comparison in our study--randomized trials showed lower viral suppression with reduced visit frequency (aRR 0.74, 95% CI 0.59-0.94) but cohort studies reported the opposite, with higher suppression among those with reduced visits (1.40, 90.95-2.08). In analysis restricted to clients with documented viral load tests, trial data showed no meaningful difference (1.00, 0.92-1.08), while among cohorts reduced visits were again associated with higher suppression (1.44, 1.24-1.66).

A number of other characteristics were associated with our main findings. Clients collecting medication from a facility-based pickup point were more likely to be not retained than those using community-based external pickup points (aRR 1.29, 95% CI 1.09–1.52), suggesting that facility pickup points offer fewer of the convenience benefits that community-based collection provides(18). This finding is particularly relevant given the recent national rollout of facility-based 6MMD, where clients collect all their medication at the facility and may not benefit from the same convenient advantages offered by community-based collection. Male clients, younger clients, and older clients were more likely to be virally unsuppressed, and men less likely to be retained, regardless of prescription length. These age and sex differences are a persistent finding across many studies that 12-month prescribing alone will not resolve(19–21). Extended prescriptions are an important step forward, but improving outcomes for men and younger and older clients will require further intervention.

In sensitivity analysis, when clients with missing viral load results were reclassified as virally unsuppressed the direction of the prescription length finding changed: clients in the 12-month group were more likely to be classified as virally unsuppressed, potentially reflecting the substantially higher proportion of missing viral load results in the 12-month group. This result points to a broader limitation of reduced visit frequency: less frequent clinical contact may also mean fewer opportunities for routine monitoring, even where treatment outcomes themselves are not compromised. Whether routine monitoring has value in itself, rather than to identify those in need of intervention, is a question for future consideration, but additional attention should be paid to ensuring that clients with 12-month scripts do receive viral load tests on schedule.

Our study has a number of limitations. Importantly, 12-month prescriptions were not randomly assigned. Clinicians made the decision about which clients received them, and it is possible that more stable, better-performing clients were preferentially offered longer scripts. We could not fully adjust for this selection process, but the broad similarity in baseline characteristics between prescription groups provides some reassurance that the two groups were not substantially different at baseline. The adjustment for ART regimen, time on ART, comorbidity status, and other clinical characteristics in the regression models also reduces, though does not eliminate, the risk of residual confounding.

Retention was defined broadly as at least one medication collection visit during the outcome assessment window of months 13 to 24. While this approach captures continued engagement, it may overestimate daily medication adherence. The high proportion of missing viral load results, particularly in the 12-month group, represents a further limitation. It was not possible to establish whether missing results reflect disengagement from care, missed testing opportunities, or documentation failures. As demonstrated in the sensitivity analysis, the direction of the viral suppression finding is sensitive to assumptions about these missing values. A related consideration is the higher proportion of clients on TLD in the 12-month group compared to the 6-month group. This most likely reflects the timing of the national dolutegravir rollout during the study period rather than selective prescribing(22), but it could have introduced an imbalance that favoured the 12-month group in the primary analysis. ART regimen was included as a covariate in all adjusted models to account for this difference.

Finally, we note that this study was conducted during the COVID-19 pandemic, and the intervention evaluated—12-month prescribing—resulted directly from pandemic concerns. Existing evidence cannot tell us whether findings can be generalised to non-pandemic periods. The enrolment period spanned varying levels of lockdown restriction, though results were broadly consistent across months, providing some reassurance that findings were not driven by a single phase of the pandemic response.

## Conclusion

This large retrospective cohort provides real-world evidence that 12-month ART prescriptions were not associated with poorer retention in care than 6-month prescriptions among stable people living with HIV enrolled in CCMDD in seven South African provinces. Twelve-month prescribing was associated with fewer clinic visits, while medication collection remained convenient through the CCMDD program. Among participants with an observed viral load result, viral suppression was similar between prescription groups, though the higher proportion of missing viral load results in the 12-month group warrants caution in interpreting the virological findings. These findings contribute to the evidence base for reduced-frequency clinical prescribing within differentiated service delivery and underscore the importance of maintaining access to routine viral-load monitoring as clinical contact is reduced.

**Supplementary Table S1.** Baseline characteristics of people living with HIV on CCMDD in South Africa between June and November 2020, disaggregated by comorbidity status (N = 428,926)?

| **Characteristic** | **HIV only<br>(N = 308,728)** | **HIV + non-HIV comorbid<br>(N = 120,198)** | **Total<br>(N = 428,926)** |
|---|---|---|---|
| **Age (years)** |  |  |  |
| *Median (IQR)* | 42 (36, 49) | 48 (42, 56) | 44 (37, 51) |
| 18–24 | 11,252 (4%) | 1,731 (2%) | 12,983 (3%) |
| 25–34 | 82,059 (28%) | 14,758 (13%) | 96,817 (24%) |
| 35–49 | 159,287 (54%) | 57,925 (51%) | 217,212 (53%) |
| 50+ | 42,097 (14%) | 38,580 (34%) | 80,677 (20%) |
| **Time on ART at baseline** |  |  |  |
| *Median (IQR)* | 6 (3, 8) | 6 (4, 9) | 6 (4, 9) |
| 6 months–1 year | 16,767 (5%) | 6,083 (5%) | 22,850 (5%) |
| 2–4 years | 99,691 (32%) | 31,179 (26%) | 130,870 (31%) |
| 5–9 years | 142,613 (46%) | 55,420 (46%) | 198,033 (46%) |
| 10+ years | 49,657 (16%) | 27,516 (23%) | 77,173 (18%) |
| **Sex** |  |  |  |
| Female | 208,456 (68%) | 83,788 (70%) | 292,244 (68%) |
| Male | 100,271 (32%) | 36,409 (30%) | 136,680 (32%) |
| Missing | 1 (0%) | 1 (0%) | 2 (0%) |
| **Prescription length (months)** |  |  |  |
| 6 months | 140,990 (46%) | 55,540 (46%) | 196,531 (46%) |
| 12 months | 167,738 (54%) | 64,658 (54%) | 232,395 (54%) |
| **Province** |  |  |  |
| Eastern Cape | 36,170 (12%) | 12,106 (10%) | 48,276 (11%) |
| Free State | 11,091 (4%) | 5,505 (5%) | 16,596 (4%) |
| Gauteng | 77,656 (25%) | 32,116 (27%) | 109,772 (26%) |
| KwaZulu-Natal | 135,453 (44%) | 40,026 (33%) | 175,479 (41%) |
| Limpopo | 4,670 (2%) | 2,810 (2%) | 7,480 (2%) |
| Mpumalanga | 30,800 (10%) | 17,219 (14%) | 48,019 (11%) |
| North West | 12,888 (4%) | 10,416 (9%) | 23,304 (5%) |
| **ART regimen** |  |  |  |
| TDF/3TC/DTG (TLD) | 167,141 (54%) | 68,398 (57%) | 235,539 (55%) |
| TDF/FTC/EFV (TEE) | 128,869 (42%) | 45,442 (38%) | 174,311 (41%) |
| Other* | 12,629 (4%) | 6,333 (5%) | 18,962 (4%) |
| Missing | 89 (0%) | 25 (0%) | 114 (0%) |
| **Model of care** |  |  |  |
| Ex-PuP (community pickup) | 197,946 (64%) | 72,692 (60%) | 270,638 (63%) |
| Fac-PuP (facility pickup) | 110,499 (36%) | 47,374 (39%) | 157,873 (37%) |
| Missing | 266 (0%) | 129 (0%) | 395 (0%) |

*IQR = interquartile range. Ex-PuP = external/community pickup point. Fac-PuP = facility pickup point. TLD = tenofovir/lamivudine/dolutegravir. TEE = tenofovir/emtricitabine/efavirenz. \*Other ART regimens comprise those prescribed following confirmed virological failure on a first-line regimen.*

**Supplementary Table S2.** Retention in care and association with characteristics of people living with HIV on CCMDD in South Africa between June and November 2020 (N = 428,926)

| Characteristic | Outcome — n (%) |  |  |  | Adjusted RR<br>(95% CI) | Unadjusted RR<br>(95% CI) |
|---|---|---|---|---|---|---|
|  | Retained<br>(N = 392,488) | Not Retained<br>(N = 36,130) | Transferred<br>(N = 214) | Died<br>(N = 94) |  |  |
| **Prescription length (months)** |  |  |  |  |  |  |
| *6 months* | 179,546 (46) | 16,736 (46) | 169 (79) | 79 (84) | 1 (ref) | 1 (ref) |
| **12 months** | 212,942 (54) | 19,394 (54) | 45 (21) | 15 (16) | 0.98 (0.91–1.05) | 0.98 (0.91–1.05) |
| **Model of care** |  |  |  |  |  |  |
| *Ex-PuP* | 250,017 (64) | 20,466 (57) | 122 (57) | 33 (35) | 1 (ref) | 1 (ref) |
| **Fac-PuP** | 142,290 (36) | 15,438 (43) | 84 (39) | 61 (65) | 1.29 (1.09–1.52) | 1.30 (1.09–1.54) |
| **Missing** | 162 (0) | 225 (1) | 8 (4) | 0 (0) | — | — |
| **Sex** |  |  |  |  |  |  |
| *Female* | 267,528 (68) | 24,499 (68) | 164 (77) | 53 (56) | 1 (ref) | 1 (ref) |
| **Male** | 124,958 (32) | 11,631 (32) | 50 (23) | 41 (44) | 1.06 (1.02–1.10) | 1.02 (0.97–1.07) |
| **Age, years** |  |  |  |  |  |  |
| *18–24* | 11,441 (3) | 1,515 (4) | 25 (12) | 2 (2) | 1 (ref) | 1 (ref) |
| **25–34** | 87,439 (23) | 9,292 (27) | 77 (37) | 9 (11) | 0.84 (0.79–0.89) | 0.81 (0.77–0.87) |
| **35–49** | 200,198 (54) | 16,908 (49) | 74 (36) | 32 (38) | 0.68 (0.63–0.74) | 0.66 (0.60–0.72) |
| **50+** | 74,055 (20) | 6,550 (19) | 31 (15) | 41 (49) | 0.70 (0.64–0.77) | 0.69 (0.629–0.76) |
| **Time on ART** |  |  |  |  |  |  |
| *6m–1yr* | 20,555 (5) | 2,274 (6) | 17 (8) | 4 (4) | 1 (ref) | 1 (ref) |
| **2–4yr** | 119,088 (30) | 11,671 (32) | 92 (43) | 19 (20) | 0.90 (0.87–0.94) | 0.90 (0.86–0.93) |
| **5–9yr** | 181,738 (46) | 16,177 (45) | 79 (37) | 39 (41) | 0.85 (0.81–0.90) | 0.82 (0.78–0.86) |
| **10yr+** | 71,107 (18) | 6,008 (17) | 26 (12) | 32 (34) | 0.84 (0.79–0.89) | 0.78 (0.74–0.83) |
| **Medication/Condition** |  |  |  |  |  |  |
| *HIV only* | 282,511 (72) | 25,991 (72) | 167 (78) | 59 (63) | 1 (ref) | 1 (ref) |
| **HIV + non-HIV** | 109,977 (28) | 10,139 (28) | 47 (22) | 35 (37) | 1.04 (0.96–1.12) | 1.00 (0.93–1.08) |
| **ART regimen** |  |  |  |  |  |  |
| *TDF/3TC/DTG (TLD)* | 216,054 (55) | 19,321 (53) | 111 (52) | 53 (56) | 1 (ref) | 1 (ref) |
| **TDF/FTC/EFV (TEE)** | 159,229 (41) | 14,959 (41) | 91 (43) | 32 (34) | 1.03 (0.99–1.07) | 1.04 (1.01–1.08) |
| **Other** | 17,113 (4) | 1,837 (5) | 6 (3) | 6 (6) | 1.23 (1.14–1.32) | 1.18 (1.10–1.26) |
| **Missing** | 92 (0) | 13 (0) | 6 (3) | 3 (3) | — | — |

*aRR = adjusted risk ratio.RR = risk ratio. CI = confidence interval. Ex-PuP = external/community-based pickup point. Fac-PuP = facility-based pickup point. TDF/3TC/DTG = tenofovir/lamivudine/dolutegravir (TLD). TDF/FTC/EFV = tenofovir/emtricitabine/efavirenz (TEE). ART = antiretroviral therapy. Reference (ref) categories shown in italics.*

**Supplementary Table S3.** Viral suppression and association with baseline characteristics of people living with HIV registered on CCMDD in South Africa between June and November 2020 and retained in care with a recorded viral load result (N = 319,319)

| **Characteristic** | **Outcome — n (%)** |  |  | **Adjusted RR<br>(95% CI)** | **Unadjusted RR<br>(95% CI)** |
|---|---|---|---|---|---|
|  | **Missing<br>(N = 109,607)** | **Virally<br>Unsuppressed<br>(N = 63,564)** | **Virally<br>Suppressed<br>(N = 255,755)** |  |  |
| **Prescription length (months)** |  |  |  |  |  |
| *6* | 42,219 (39) | 31,994 (50) | 122,317 (48) | 1 (ref) | 1 (ref) |
| **12** | 67,388 (61) | 31,570 (50) | 133,438 (52) | 0.93 (0.82–1.06) | 0.93 (0.80–1.07) |
| **Model of care** |  |  |  |  |  |
| *Ex-PuP* | 70,098 (64) | 38,539 (61) | 162,001 (63) | 1 (ref) | 1 (ref) |
| **Fac-PuP** | 39,382 (36) | 24,922 (39) | 93,569 (37) | 1.10 (0.98–1.24) | 1.10 (0.98–1.23) |
| **Missing** | 124 (0) | 100 (0) | 171 (0) | — | — |
| **Sex** |  |  |  |  |  |
| *Female* | 73,020 (67) | 39,982 (63) | 179,242 (70) | 1 (ref) | 1 (ref) |
| **Male** | 36,585 (33) | 23,582 (37) | 76,513 (30) | 1.260 (1.210–1.312) | 1.29 (1.23–1.36) |
| **Missing** | 2 (0) | 0 (0) | 0 (0) | — | — |
| **Age group** |  |  |  |  |  |
| *18–24* | 3,602 (3) | 1,571 (3) | 7,810 (3) | 1 (ref) | 1 (ref) |
| **25–34** | 25,216 (24) | 12,137 (20) | 59,464 (24) | 1.01 (0.96–1.05) | 1.019 (0.97–1.07) |
| **35–49** | 54,916 (53) | 33,093 (55) | 129,203 (53) | 1.14 (1.07–1.21) | 1.23 (1.15–1.32) |
| **50+** | 20,753 (20) | 13,003 (22) | 46,921 (19) | 1.17 (1.09–1.27) | 1.32 (1.22–1.43) |
| **Time on ART** |  |  |  |  |  |
| *6m–1yr* | 7,030 (6) | 3,151 (5) | 12,669 (5) | 1 (ref) | 1 (ref) |
| **2–4yr** | 34,429 (31) | 17,949 (28) | 78,492 (31) | 0.92 (0.89–0.96) | 0.94 (0.90–0.98) |
| **5–9yr** | 49,248 (45) | 30,126 (47) | 118,659 (46) | 0.97 (0.92–1.03) | 1.01 (0.96–1.09) |
| **10yr+** | 18,900 (17) | 12,338 (19) | 45,935 (18) | 0.99 (0.91–1.07) | 1.07 (0.97–1.17) |
| **Condition** |  |  |  |  |  |
| *HIV only* | 79,695 (73) | 44,512 (70) | 184,521 (72) | 1 (ref) | 1 (ref) |
| **HIV + non-HIV** | 29,912 (27) | 19,052 (30) | 71,234 (28) | 1.05 (0.99–1.11) | 1.10 (1.02–1.17) |
| **ART regimen** |  |  |  |  |  |
| *TDF/3TC/DTG* | 60,419 (55) | 35,407 (56) | 139,713 (55) | 1 (ref) | 1 (ref) |
| **TDF/FTC/EFV** | 44,284 (40) | 24,267 (38) | 105,760 (41) | 0.96 (0.90–1.03) | 0.921 (0.86–0.98) |
| **Other** | 4,801 (4) | 3,887 (6) | 10,274 (4) | 1.36 (1.25–1.48) | 1.34 (1.24–1.45) |
| **Missing** | 103 (0) | 3 (0) | 8 (0) | — | — |

*aRR = adjusted risk ratio. Adjusted for: prescription length, model of care, sex, age group, time on ART, condition, ART regimen, and province. The 95% confidence interval represents the range of values within which the true risk ratio is likely to fall. Narrower intervals indicate more precise estimates.— = not modelled (descriptive only).*

**Supplementary Table S4.**
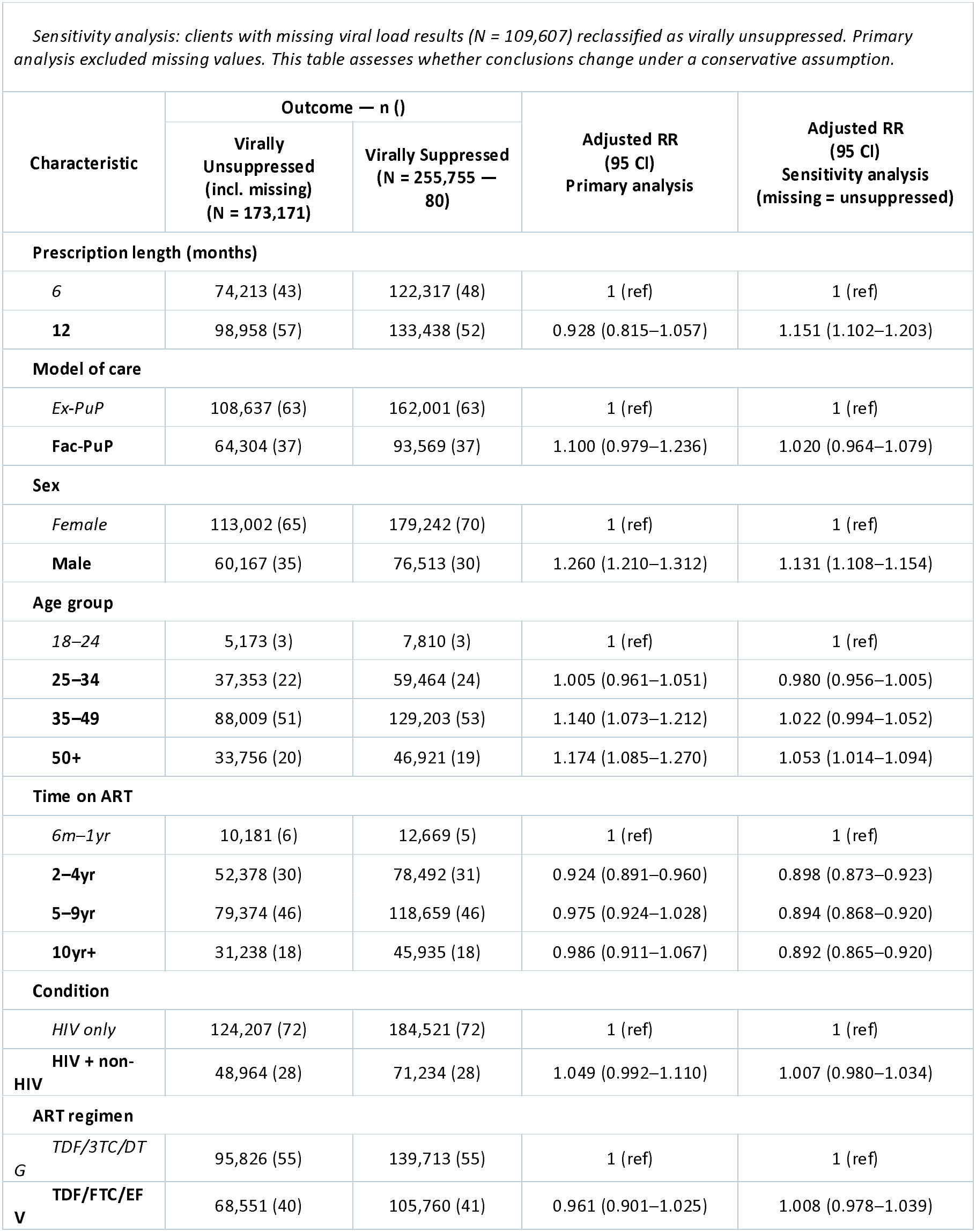
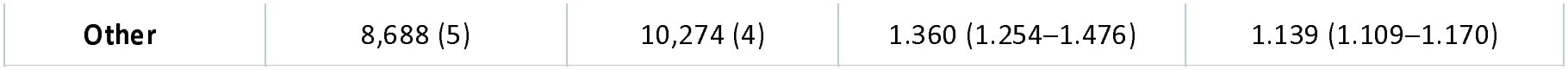
Sensitivity analysis — viral suppression treating missing viral load results as virally unsuppressed, among people living with HIV registered on CCMDD in South Africa between June and November 2020 and retained in care with a recorded viral load result (N = 319,319) (N = 319,319)

| Characteristic | Outcome — n () |  | Adjusted RR<br>(95 CI)<br>Primary analysis | Adjusted RR<br>(95 CI)<br>Sensitivity analysis<br>(missing = unsuppressed) |
|---|---|---|---|---|
|  | Virally Unsuppressed (incl. missing) (N = 173,171) | Virally Suppressed (N = 255,755 — 80) |  |  |
| **Prescription length (months)** |  |  |  |  |
| *6* | 74,213 (43) | 122,317 (48) | 1 (ref) | 1 (ref) |
| **12** | 98,958 (57) | 133,438 (52) | 0.928 (0.815–1.057) | 1.151 (1.102–1.203) |
| **Model of care** |  |  |  |  |
| *Ex-PuP* | 108,637 (63) | 162,001 (63) | 1 (ref) | 1 (ref) |
| **Fac-PuP** | 64,304 (37) | 93,569 (37) | 1.100 (0.979–1.236) | 1.020 (0.964–1.079) |
| **Sex** |  |  |  |  |
| *Female* | 113,002 (65) | 179,242 (70) | 1 (ref) | 1 (ref) |
| **Male** | 60,167 (35) | 76,513 (30) | 1.260 (1.210–1.312) | 1.131 (1.108–1.154) |
| **Age group** |  |  |  |  |
| *18–24* | 5,173 (3) | 7,810 (3) | 1 (ref) | 1 (ref) |
| **25–34** | 37,353 (22) | 59,464 (24) | 1.005 (0.961–1.051) | 0.980 (0.956–1.005) |
| **35–49** | 88,009 (51) | 129,203 (53) | 1.140 (1.073–1.212) | 1.022 (0.994–1.052) |
| **50+** | 33,756 (20) | 46,921 (19) | 1.174 (1.085–1.270) | 1.053 (1.014–1.094) |
| **Time on ART** |  |  |  |  |
| *6m–1yr* | 10,181 (6) | 12,669 (5) | 1 (ref) | 1 (ref) |
| **2–4yr** | 52,378 (30) | 78,492 (31) | 0.924 (0.891–0.960) | 0.898 (0.873–0.923) |
| **5–9yr** | 79,374 (46) | 118,659 (46) | 0.975 (0.924–1.028) | 0.894 (0.868–0.920) |
| **10yr+** | 31,238 (18) | 45,935 (18) | 0.986 (0.911–1.067) | 0.892 (0.865–0.920) |
| **Condition** |  |  |  |  |
| *HIV only* | 124,207 (72) | 184,521 (72) | 1 (ref) | 1 (ref) |
| **HIV + non-HIV** | 48,964 (28) | 71,234 (28) | 1.049 (0.992–1.110) | 1.007 (0.980–1.034) |
| **ART regimen** |  |  |  |  |
| *TDF/3TC/DTG* | 95,826 (55) | 139,713 (55) | 1 (ref) | 1 (ref) |
| **TDF/FTC/EFV** | 68,551 (40) | 105,760 (41) | 0.961 (0.901–1.025) | 1.008 (0.978–1.039) |
| **Other** | 8,688 (5) | 10,274 (4) | 1.360 (1.254–1.476) | 1.139 (1.109–1.170) |

## Data Availability

All data used in this study were collected by the study team following written informed consent. Data will be made publicly available in a data repository within one year of study closure, as approved by the supervising ethics committees. The corresponding author can be contacted for details.

## References

1. Spotlight. SA’s ARV programme hardly grew in 2025, according to latest estimate [Internet]. 2026 [cited 2026 Aug 21]. Available from: https://www.spotlightnsp.co.za/2026/05/15/sas-arv-programme-hardly-grew-in-2025-according-to-latest-estimates/

2. World Health Organisation. Consolidated guidelines on HIV prevention, testing, treatment, service delivery and monitoring⍰: recommendations for a public health approach [Internet]. Geneva, editor. World Health Organization; 2021 [cited 2025 Jul 21]. 548 p. Available from: https://www.who.int/publications/i/item/9789240031593

3. Schwartz SR, Baral S. Remembering individual perspectives and needs in differentiated HIV care strategies. BMJ Quality and Safety. BMJ Publishing Group; 2019. p. 257–9. doi:10.1136/bmjqs-2018-008339 PubMed PMID: 30591539.

4. South African National Department of Health. National Consolidated Guidelines For the Management of HIV in Adults, Adolescents, Children and Infants and Prevention of Vertical Transmission [Internet]. 2026 [cited 2026 Jul 17]. Available from: https://sahivsoc.org/Files/National%20Consolidated%20Guidelines%2004-03-2026.pdf

5. South African National Department of Health: Government Notice. Medicines and Related Substances Act: Exclusion of Schedule 2, Schedule 3 and Schedule 4 substances from the operation of certain provisions [Internet]. 2020. Available from: http://www.gpwonline.co.za

6. Tourneau N Le, German A, Thompson R, Ford N, Shwartz S, Beres L, et al. Evaluation of HIV treatment outcomes with reduced frequency of clinical encounters and antiretroviral treatment refills: A systematic review and meta-analysis. PLoS Medicine. Public Library of Science; 2022. doi:10.1371/journal.pmed.1003959 PubMed PMID: 35316272.

7. South African National Department of Health. National consolidated guidelines for the management of HIV in adults, adolescents, children and infants and prevention of mother-to-child transmission [Internet]. 2020 [cited 2024 Sep 6]. Available from: https://knowledgehub.health.gov.za/system/files/elibdownloads/2023-04/National%2520Consolidated%2520Guidelines%252030062020%2520signed%2520PRINT%2520v7.pdf

8. National Department of Health. Minimum differentiated models of care packages to support linkage of care, adherence and retention in care. Differentiated models of care: Standard operating procedures and adherence guidelines for HIV, TB and NCDs. 2023.

9. Liu L, Christie S, Munsamy M, Roberts P, Pillay M, Shenoi S V., et al. Title: Expansion of a national differentiated service delivery model to support people living with HIV and other chronic conditions in South Africa: a descriptive analysis. BMC Health Serv Res. 2021 Dec 1;21(1). doi:10.1186/s12913-021-06450-z PubMed PMID: 34001123.

10. Bogart LM, Shazi Z, MacCarthy S, Mendoza-Graf A, Wara NJ, Zionts D, et al. Implementation of South Africa’s Central Chronic Medicine Dispensing and Distribution Program for HIV Treatment: A Qualitative Evaluation. AIDS Behav. 2022 Aug 1;26(8):2600–12. doi:10.1007/s10461-022-03602-y PubMed PMID: 35122215.

11. Tlhaku K, Msimango L, Sookrajh Y, Milford C, Munatsi P, Gray A, et al. Healthcare worker perspectives on adaptations to differentiated anti-retroviral therapy delivery during COVID-19 in South Africa: A qualitative inquiry. PLOS Global Public Health. 2024 Aug 9;4(8). doi:10.1371/journal.pgph.0003517

12. Naidu L, van der Molen J, Jugathpal V, Sookrajh Y, Khubone T, Hobe L, et al. Quality assessment of data for decentralised antiretroviral therapy referrals and laboratory results in the South African national electronic HIV management register TIER.Net. Henao-Martínez AF, editor. PLOS Global Public Health. 2026 Mar 16;6(3):e0005534. doi:10.1371/journal.pgph.0005534

13. National Department of Health: Republic of South Africa. Synchronised National Communication in Health (SyNCH) [Internet]. 2023 [cited 2024 Sep 6]. Available from: https://www.hst.org.za/Documents/SyNCH%20A5%20Flyer%202023.pdf

14. Lewis L, Sookrajh Y, Van Der Molen J, Khubone T, Sosibo P, Maraj M, et al. Clinical outcomes after extended 12-month antiretroviral therapy prescriptions in a community-based differentiated HIV service delivery programme in South Africa: a retrospective cohort study. J Int AIDS Soc. 2023;26:26164. doi:10.1002/jia2.26164/full

15. Ainembabazi B, Ssebunya RN, Akobye W, Mugume A, Nahirya-Ntege P, Birungi DJ, et al. Viral load suppression and retention in care among children and adolescents receiving multi-month anti-retroviral therapy refills: a program data review in Uganda. BMC Pediatr. 2024 Dec 1;24(1). doi:10.1186/s12887-024-05295-9 PubMed PMID: 39645566.

16. Jo Y, Jamieson L, Phiri B, Grimsrud A, Mwansa M, Shakwelele H, et al. Attrition from HIV treatment after enrollment in a differentiated service delivery model: A cohort analysis of routine care in Zambia. PLoS One. 2023 Mar 1;18(3 March). doi:10.1371/journal.pone.0280748 PubMed PMID: 36917568.

17. Fatti G, Ngorima-Mabhena N, Mothibi E, Muzenda T, Choto R, Kasu T, et al. Outcomes of Three-Versus Six-Monthly Dispensing of Antiretroviral Treatment (ART) for Stable HIV Patients in Community ART Refill Groups: A Cluster-Randomized Trial in Zimbabwe [Internet]. 2020. Available from: www.jaids.com

18. Bassett I V., Yan J, Govere S, Khumalo A, Ngobese N, Shazi Z, et al. Uptake of community-versus clinic-based antiretroviral therapy dispensing in the Central Chronic Medication Dispensing and Distribution program in South Africa. J Int AIDS Soc. 2022 Jan 1;25(1):e25877. doi:10.1002/JIA2.25877 PubMed PMID: 35077611.

19. Jamieson L, Rosen S, Meyer-Rath G, Mokhele I, Musakwa N, Imai-Eaton JW, et al. Age, sex- and what else? Rethinking priorities to close gaps in the HIV care cascade. J Int AIDS Soc. 2026;29:70080. doi:10.1002/jia2.70080/full

20. Mukumbang FC. Leaving no man behind: How differentiated service delivery models increase men’s engagement in HIV care. Int J Health Policy Manag. 2021 Mar 1;10(3):129–40. doi:10.34172/ijhpm.2020.32 PubMed PMID: 32610748.

21. Chikwari CD, Kranzer K, Simms V, Patel A, Tembo M, Mugurungi O, et al. Differentiated care for youth in Zimbabwe: Outcomes across the HIV care cascade. PLOS Global Public Health. 2024 Feb 1;4(2 February). doi:10.1371/journal.pgph.0002553

22. Republic of South Africa National Department of Health. ART Clinical Guidelines for the Management of HIV in Adults, Pregnancy, Adolescents, Children, Infants and Neonates. 2019.

